# JOINT CLINICAL AND MOLECULAR SUBTYPING OF COPD WITH VARIATIONAL AUTOENCODERS

**DOI:** 10.1101/2023.08.19.23294298

**Authors:** Enrico Maiorino, Margherita De Marzio, Zhonghui Xu, Jeong H. Yun, Robert P. Chase, Craig P. Hersh, Scott T. Weiss, Edwin K. Silverman, Peter J. Castaldi, Kimberly Glass

## Abstract

Chronic Obstructive Pulmonary Disease (COPD) is a complex, heterogeneous disease. Traditional subtyping methods generally focus on either the clinical manifestations or the molecular endotypes of the disease, resulting in classifications that do not fully capture the disease’s complexity. Here, we bridge this gap by introducing a subtyping pipeline that integrates clinical and gene expression data with variational autoencoders. We apply this methodology to the COPDGene study, a large study of current and former smoking individuals with and without COPD. Our approach generates a set of vector embeddings, called Personalized Integrated Profiles (PIPs), that recapitulate the joint clinical and molecular state of the subjects in the study. Prediction experiments show that the PIPs have a predictive accuracy comparable to or better than other embedding approaches. Using trajectory learning approaches, we analyze the main trajectories of variation in the PIP space and identify five well-separated subtypes with distinct clinical phenotypes, expression signatures, and disease outcomes. Notably, these subtypes are more robust to data resampling compared to those identified using traditional clustering approaches. Overall, our findings provide new avenues to establish fine-grained associations between the clinical characteristics, molecular processes, and disease outcomes of COPD.

## 1 Introduction

Chronic Obstructive Pulmonary Disease (COPD) is a complex chronic respiratory disease that is among the leading causes of death worldwide [1]. The disease manifests as a spectrum of conditions, including persistent airflow obstruction, lung inflammation, chronic bronchitis, and emphysema. COPD susceptibility has been attributed to a combination of physical, environmental, and genetic factors, resulting in significant phenotypic variation across individuals.

This heterogeneity has prompted substantial research efforts aimed at dissecting the various manifestations of the disease, to understand their etiological origins, and to predict their outcomes [2]. A practical goal of these studies has been to delineate distinct COPD subtypes by employing advanced clustering and machine learning techniques trained with large datasets of clinical and genomic data extracted from human cohort studies [3, 4].

Current COPD subtyping approaches can be divided into those that characterize the observable phenotypes of the disease (clinical subtyping), and those that focus on disentangling the disease processes, often referred to as “endotypes”, underlying various COPD manifestations (molecular subtyping) [5]. Applications of the former type leverage clinical data including demographics, disease symptoms, spirometry measurements, or chest imaging [6, 7, 8, 9], whereas applications of the latter type leverage measurements from various omics assays (transcriptomics, proteomics, epigenomics, etc.) [10, 11, 5, 12]. Although both approaches offer valuable insights into different aspects of the disease, subtype classifications from these applications are exclusively defined within either the clinical or molecular domain, with the analysis in the other domain performed primarily for validation or post-hoc examination [13, 6, 8, 11, 5]. Consequently, these domain-specific classifications cannot capture disease mechanisms arising from the interaction between molecular processes and clinical or lifestyle factors [14, 15, 16, 17], potentially leading to inconsistent subtypes that are not reproducible across different patient cohorts [18].

Multi-omics data integration is a widely researched subject [19, 20, 21], and multiple methods have been developed for this purpose, such as MOFA [22], iCluster [23] and SNF [24]. In contrast, the simultaneous integration of both clinical and omics data for disease subtyping has received comparatively less attention, with its applications in COPD being confined to specific domains [25]. One of the challenges in integrating omics and non-omics data is their inherent complexity. Data heterogeneity and bias, already present in multi-omics studies [26], are exacerbated when including clinical data, which is typically composed of complex data structures with heterogeneous correlation patterns and significant variation in terms of scales, sparsity, and noise [27]. To alleviate these issues and to account for potential nonlinear interactions between variables from different domains, specialized integrative methodologies based on autoencoder neural networks have been proposed in several disease contexts, including COPD [10] and cancer [28, 29, 30, 31]. However, a comprehensive subtyping analysis that integrates both the clinical and molecular domains of COPD has not been performed.

In this work, we propose a joint subtyping approach to integrate clinical and gene expression data extracted from the COPDGene Study [32] (see Fig. 1 (a)), a large study of current and former smokers with and without COPD. Building on recent developments in multi-modal learning [33, 34], we developed an integrative method based on variational autoencoders (VAEs). A VAE is an unsupervised neural network architecture designed to compress the input data and generate a set of compact encodings [35]. We trained the VAE with clinical, imaging, and transcriptomic data from COPDGene, generating a set of personalized integrated profiles (PIPs) that encode the joint clinical and molecular configuration of every individual in the population. By performing multiple outcome prediction experiments, we demonstrate that the generated PIPs are highly informative of the individual’s disease state and enable accurate prediction of future disease outcomes. Next, we map the continuous trajectories in the VAE space using a recently proposed trajectory learning technique [36]. Through this approach, we identify several well-separated disease states, each exhibiting distinct clinical and molecular characteristics (joint subtypes). Finally, we show that these joint subtypes are characterized by different disease progression patterns and mortality and that they are robust to resampling noise.

**Figure 1:**
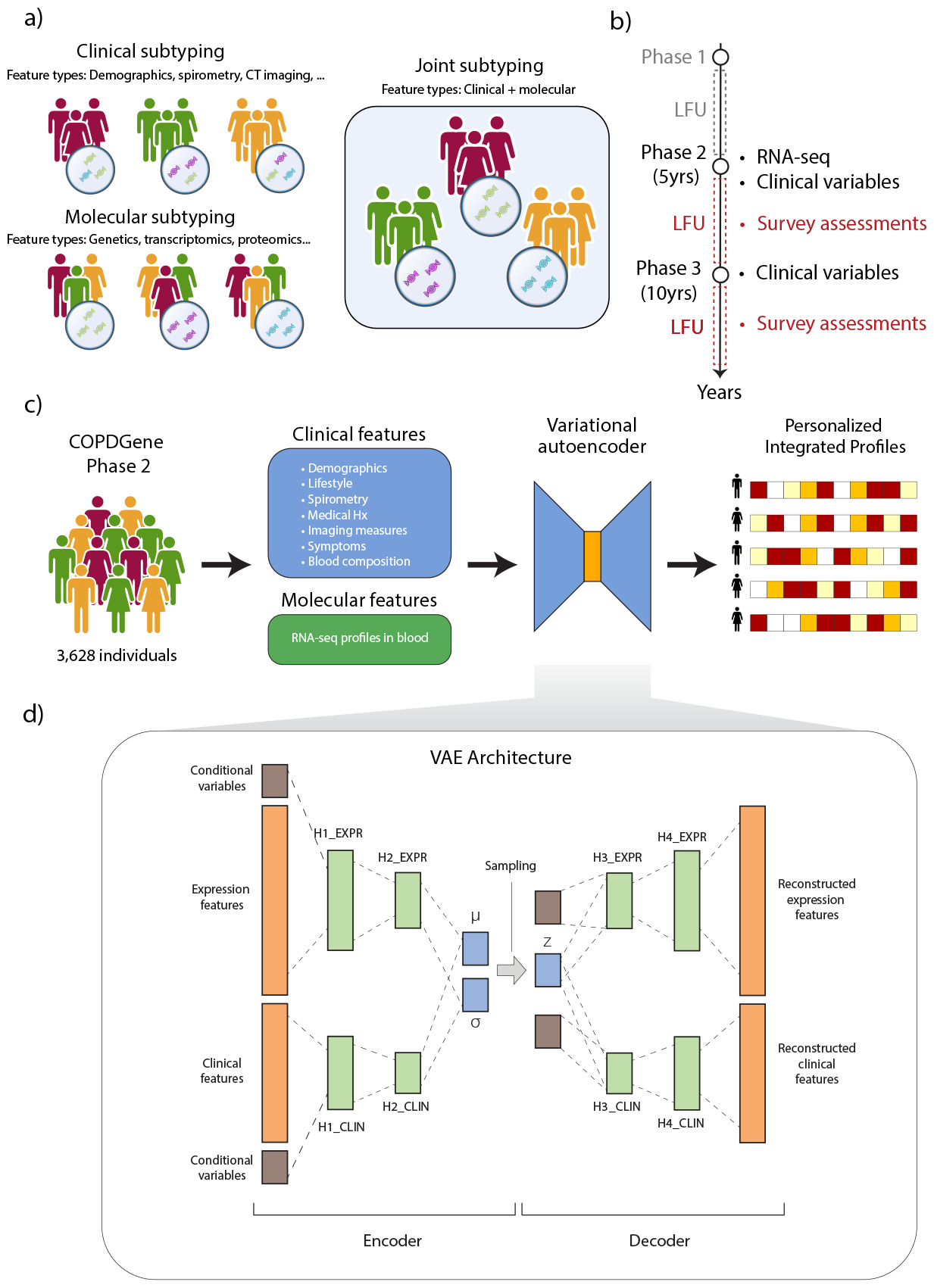
Overall organization of this work. (a) Joint subtyping aims at generating subtypes based on both clinical and molecular features. (b) COPDGene study design. Data from study stages preceding Phase 2 (gray) are not used in this work. On the right of the timeline are described the types of data collected at each study stage and considered in this study. Phase 2 and Phase 3 of the study are spaced approximately 5 years apart. LFU (Long-term Follow-Up) data consists of survey assessments conducted approximately every 6 months in between study phases. (c) Starting from the COPDGene population, an array of clinical and molecular features is extracted and used for training the VAE. The autoencoder produces a set of personalized integrated profiles (PIPs), one per individual in the population. (d) Architecture of the VAE. Expression and clinical features are concatenated with conditional variables (age, sex, race) and processed separately in two encoder subnetworks. The two encodings are subsequently merged in a shared latent representation (PIP). Next, the latent representation is concatenated to the conditional variables and processed with two separate decoder subnetworks to reconstruct the original data.

## 2 Results

COPDGene is an ongoing longitudinal multi-center study of current or ex-smoking individuals with and without COPD who have undergone extensive clinical, physiological, and radiological profiling at three time points across 10 years (Phase 1, 2, 3, see Fig. 1 (b)). Additionally, periodic long-term follow-up (LFU) surveys have been conducted every 6 months throughout the duration of the study. 10,198 individuals were enrolled at baseline. In this work we considered all of the subjects with clinical and blood gene expression data in Phase 2 (five-year follow-up) of the study that were available at the time of analysis (3,628 subjects, see Fig. 1 (c)). Clinical data includes demographics, lifestyle factors (e.g., smoking habits), spirometry measurements, medical and medication history, chest CT imaging measures, respiratory symptoms, and complete blood counts (CBC). Gene expression data consists of whole blood RNA-seq profiling. Both of these data modalities have been used extensively for COPD subtyping [6, 7, 8, 9, 11, 5], and they are among the most widely-used read-outs of the phenotypic and molecular manifestations of COPD. To merge these two data types in a single representation of disease state, we designed a data integration scheme based on Variational Autoencoders (VAEs).

VAEs are probabilistic unsupervised neural network models designed to compress the input to generate low dimensional representations [37]. VAEs exploit statistical dependencies between input variables to construct a small set of variables (latent code) that preserve most of the input information. In contrast to linear techniques such as Principal Component Analysis (PCA), VAEs can capture nonlinear relationships between variables [38]. Drawing inspiration from other recently proposed architectures for multi-modal data integration [29, 30], we modified the standard VAE architecture so as to process and merge two data types (see Fig. 1 (d) and Methods). Our model implicitly performs a 2-step dimensionality reduction. The hidden layers in the encoder network (H1/2-expr, H1/2-clin in Fig. 1 (d)) process the two data types separately to obtain a data-type-specific representation of the input features. These representations are then coupled to generate a joint latent representation that encodes both the clinical and molecular information, which we refer to as “Personalized Integrated Profile” (PIP). Given the probabilistic nature of the VAE model, it is possible to correct for potential confounding factors by including them as a set of conditional variables [39]. These conditional variables have a similar role as regression covariates in linear regression modeling. Therefore, we set the age, sex, and race of each individual as conditional variables in both data modalities to regress out their effect on the learned representation.

The data processing and integration pipeline consists of several steps (see Methods for further details). In brief, we designed two separate processing pipelines for the two data types, consisting of feature selection and normalization operations. The resulting set of features selected for training the VAE is summarized in Supplementary Table 1. Next, we split the dataset into training and validation sets (80%/20%). We trained the VAE on the training set, and used the validation set for hyperparameter selection. We performed hyperparameter optimization to determine the optimal network depth, layer size, learning rate, and number of components of PIP vectors (latent components). Our primary objective was to select parameter configurations that resulted in high reconstruction accuracy of the validation set data, with a preference for configurations with fewer latent components. We determined that the optimal number of components in terms of reconstruction quality and latent vector size is 30. After training the network with the optimal parameters, we then used the Encoder subnetwork to generate the PIPs of the full dataset. The generated PIPs are the starting point of the subsequent analysis to identify joint subtypes of COPD.

### 2.1 Predicting future disease states with Personalized Integrated Profiles

The PIPs generated by the VAE contain information on both the expression and clinical features of a subject. To test the VAE’s performance in compressing and integrating different data modalities without sacrificing important information, we set up a prediction task of several prospective COPD outcomes. The COPD outcomes included all-cause mortality at 3 and 5 years after the Phase 2 and other clinical measurements collected in Phase 3 (P3) of the study, approximately five years after Phase 2 (P2) visits (see Methods for further details). These variables are extracted exclusively from data collected after Phase 2 of the study and thus were not used to train the VAE. We perform the classification using a Random Forest classifier, and define the PIP vectors as input variables and the Phase 3 outcomes as target variables (see Methods for details). For comparison, we evaluated the performance of the same classifier trained using other types of embedding as input: PCA of clinical variables (Clin PCA); PCA of expression variables (Expr PCA); PCA of concatenated expression and clinical variables (Expr + Clin PCA); Canonical Correlation Analysis (CCA) scores of the expression (Expr CCA) and clinical variables (Clin CCA); and factors calculated by applying the integrative method Multi Omics Factor Analysis (MOFA) [22] to expression and clinical variables. The performance metrics for the prediction, presented in Table 1, were derived from a 5-fold stratified validation repeated 3 times and then averaged (additional measures are reported in Supplementary Table 2). The VAE-based PIPs consistently achieve either the highest or second-highest scores across most prediction tasks. In the cases where the PIPs do not outperform other embeddings, no other alternative embedding scheme emerges as a clear leader in performance. This finding indicates that despite encoding a broader range of information compared to domain-specific alternatives (e.g. Clin PCA), the PIPs retain substantial information about an individual’s disease state and its likely outcomes.

**Table 1:** Prediction performance (F1-score) of COPD outcomes. Best performances and second-best are respectively displayed in bold or underlined. Abbreviations: Δ = change from P2 to P3, P2 = phase 2 of COPDGene, P3 = phase 3 of COPDGene, (P3>P2) = value at P3 larger than value at P2, pred.=predicted, inc. chronic bronchitis = chronic bronchitis not present at P2 but present at P3, exac. freq.=exacerbations frequency

|  | F1-score |  |  |  |  |  |  |
| --- | --- | --- | --- | --- | --- | --- | --- |
|  | Clin PCA | Expr PCA | Expr+Clin PCA | Expr CCA | Clin CCA | MOFA | VAE |
| $\Delta$ FEV<sub>1</sub> % of pred. | <u>0.75 (0.03)</u> | 0.74 (0.04) | 0.73 (0.03) | **0.75 (0.04)** [n.s.] | 0.74 (0.04) | 0.74 (0.04) | 0.73 (0.04) |
| Inc. chronic bronchitis | <u>0.02 (0.04)</u> | 0.01 (0.03) | 0.04 (0.06) | 0.03 (0.05) | 0.04 (0.05) | <u>0.07 (0.07)</u> | **0.12 (0.08)** ** |
| Exacerbations (P3) | <u>0.37 (0.09)</u> | 0.09 (0.06) | 0.35 (0.06) | 0.23 (0.07) | 0.23 (0.08) | <u>0.25 (0.09)</u> | **0.41 (0.08)** * |
| $\Delta$ Exac. Freq. (P3>P2) | <u>0.10 (0.08)</u> | 0.04 (0.05) | 0.14 (0.07) | 0.12 (0.07) | 0.12 (0.08) | 0.09 (0.06) | **0.20 (0.11)** ** |
| Sev. Exacerbations (P3) | <u>0.11 (0.07)</u> | 0.04 (0.04) | 0.11 (0.08) | 0.04 (0.05) | 0.03 (0.04) | 0.06 (0.06) | **0.15 (0.08)** * |
| $\Delta$ Sev. Exacerbations (P3>P2) | <u>0.08 (0.06)</u> | 0.03 (0.05) | 0.06 (0.08) | 0.00 (0.02) | 0.01 (0.03) | 0.04 (0.06) | **0.11 (0.07)** [n.s.] |
| $\Delta$ MMRC (P3>P2) | **0.29 (0.05)** * | 0.18 (0.05) | 0.18 (0.05) | 0.24 (0.04) | 0.22 (0.05) | 0.20 (0.05) | 0.26 (0.04) |
| $\Delta$ SF-36 (P3<P2) | 0.58 (0.03) | 0.59 (0.03) | 0.60 (0.03) | <u>0.60 (0.02)</u> | **0.61 (0.02)** * | 0.58 (0.03) | 0.59 (0.03) |
| Mortality (3yr) | 0.08 (0.04) | 0.12 (0.05) | 0.11 (0.05) | <u>0.13 (0.06)</u> | 0.13 (0.06) | 0.10 (0.03) | **0.17 (0.05)** * |
| Mortality (5yr) | 0.23 (0.05) | 0.19 (0.04) | 0.23 (0.03) | 0.25 (0.05) | <u>0.25 (0.04)</u> | 0.20 (0.04) | **0.30 (0.06)** ** |

### 2.2 Summarizing COPD heterogeneity with principal graphs

The PIPs generated by the VAE are vectors distributed in a 30-dimensional space of variables that implicitly describe the joint molecular and clinical characterization of every individual analyzed in our COPDGene dataset. The geometry of the distribution of these generated vectors in the VAE space is therefore informative of the patterns of variability of COPD features in the population, including the presence, or lack thereof, of separate clusters. Growing evidence suggests that COPD manifestations may form a continuous spectrum of disease states[3]. Under such circumstances, characterizations based on discrete clusters may impose arbitrary boundaries within subpopulations that may impact the robustness of the subtypes. To overcome this issue, we analyzed the trajectories of continuous variation of COPD in the VAE space. We used the elPiGraph method [36], an algorithm to fit a branching network structure to a set of points in a multidimensional space. ElPiGraph and its adaptations have been previously used to identify trajectories in several contexts, including the clinical domain of COPD [40] and the molecular domain of cancer [41]. In this study, we extend its application to the joint domain of clinical and molecular features of COPD.

In brief, elPiGraph produces a tree-like network, called *principal graph*, that is embedded in the VAE space. elPiGraph optimizes the coordinates of the nodes of the principal graph to minimize their distance from the data points. In this way, the principal graph traverses the main axes of variation of the data points, approximating their intrinsic geometry. This procedure results in a mapping between each data point and its corresponding projection onto the tree branches, providing information on their relative positioning along the main axes of variation.

We applied elPiGraph to construct the principal graph of the population and associated each subject to their closest network branch in the space. The fitted principal graph is composed of 5 terminal branches, i.e., tree branches connected to the remaining graph only through one endpoint, and 2 non-terminal branches, i.e., those connected at both their endpoints (see Fig. 2 (a)). Since we were interested in identifying subtypes with distinct disease features and minimal overlap, we restricted the analysis exclusively to the individuals within the terminal branches. Further, to maximize the separation between different branches, we selected only the 50% of data points that lie on the most extreme ends of each branch (see Methods and Figs. 2 (b) and Supplementary Figs. 1 (a-f)). Overall, 1,552 individuals were selected as members of any of the 5 branches.

**Figure 2:**
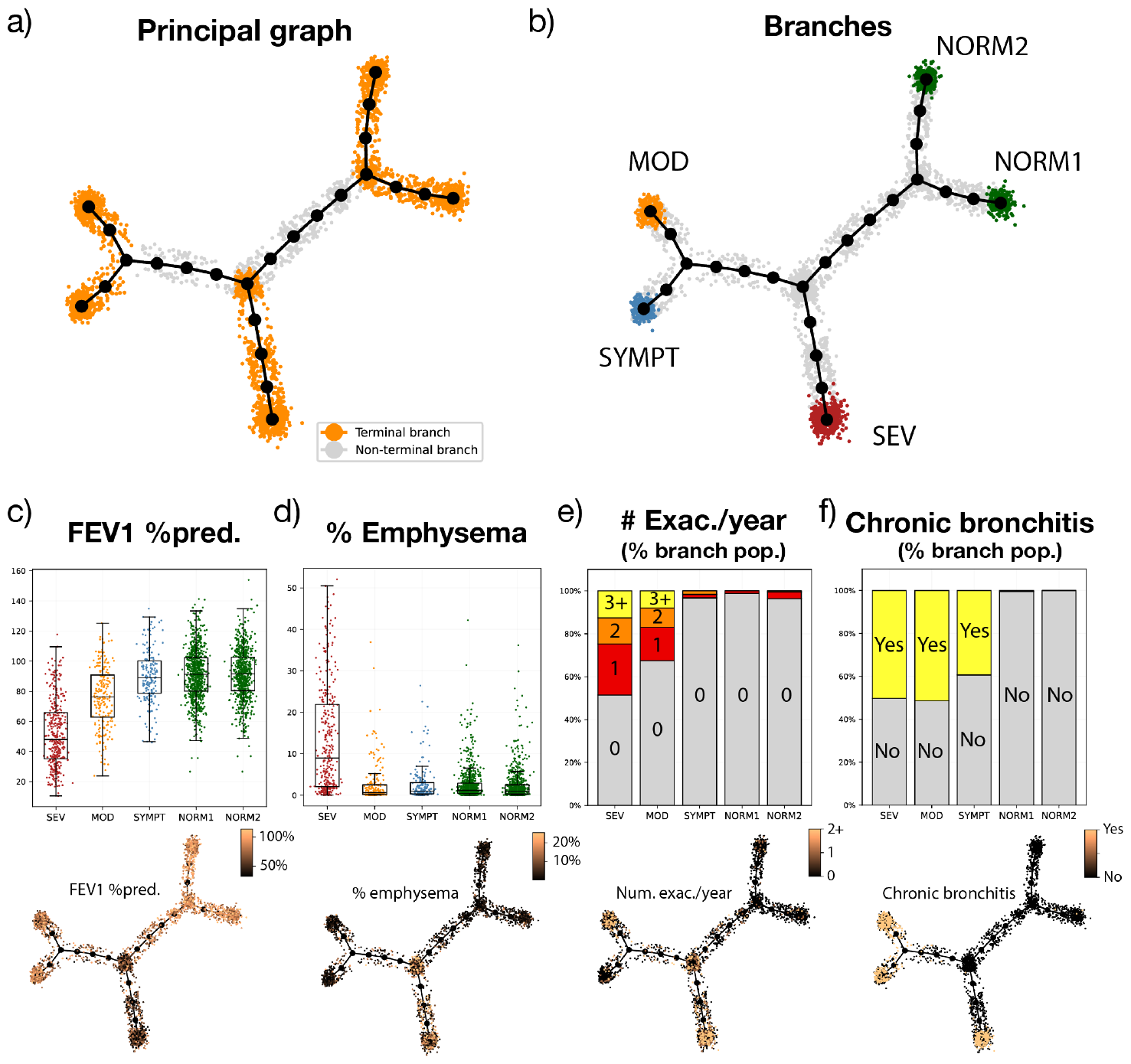
Features of the principal graph constructed from the COPDGene PIPs. (a) Principal graph layout. Large black dots and edges represent the fitted graph structure. Smaller dots represent individuals in the population, where the position is determined according to their proximity to the graph nodes. Points highlighted in orange are members of the terminal branches, while gray points are in non-terminal branches. (b) Colored dots represent the individuals selected as members of each branch after thresholding. Gray dots are individuals that are not assigned to any branch. (c-f) Distribution of selected features in each branch (top) and across the principal graph (bottom). All the graph layouts are generated with the Kamada-Kawai algorithm. Abbreviations: Pred.=predicted, Exac.=Exacerbations

### 2.3 The joint subtypes have distinct clinical characteristics

Of the 5 terminal branches, 3 are composed mainly of individuals with GOLD stage 2-4 corresponding to moderate-to-severe COPD (bottom left branches in Fig. 2 (b)), whereas 2 branches (referred to as NORM1 and NORM2) are composed mainly of individuals with preserved lung function or mild COPD, i.e. GOLD stage equal to 0-1 (top right branches in Fig. 2 (b)). The branches are characterized by different phenotypic profiles (see Supplementary Table 3).

The NORM1 and NORM2 branches differ primarily in mean age (NORM1: 67±8.2 yrs, NORM2: 61 ± 8.5 yrs), current smoking status (NORM1: 18%, NORM2: 44%), and blood cell type composition (predominantly neutrophilic in NORM1 and lymphocytic in NORM2).

The SEV branch consists of older individuals (68.7 ± 8.3 yrs) with the most severe disease manifestations - low lung function (52% of predicted FEV_1_), frequent and severe exacerbations, and high prevalence of chronic bronchitis (Figs. 2 (c-f)). These individuals also have a marked degree of emphysema, air trapping, and thickened airway walls. Consistent with the severity of their condition, self-reported metrics such as the mMRC dyspnoea scale and SGRQ scores indicate a severely compromised quality of life. Despite the high average number of pack years, the SEV branch is characterized by the smallest proportion of current smokers (31%) compared to other branches. This trend likely reflects the tendency of subjects with severe COPD to stop smoking.

The MOD branch consists of younger individuals (59.7±6.3 yrs) with moderately impaired lung function and low percentage of emphysema and moderate airway wall thickening. However, compared to other branches (excluding SEV), MOD subjects are affected by a higher average frequency of exacerbations, and the severity of their respiratory symptoms is similar to those observed in the SEV branch, with an average mMRC score of 2.4±1.3 compared to 2.7±1.2 in the SEV branch and an SGRQ total score of 46.2±19.7 versus 48.7±18.6. In line with previous studies [42], we defined frequent exacerbators as those individuals who experience two or more exacerbations in a year. The MOD branch has the second-largest proportion of frequent exacerbators (17%) after SEV (25%), and this subgroup has a substantial proportion of subjects in GOLD spirometric stage 2 or PRISm (preserved ratio impaired spirometry) group [43] (Supplementary Figure 2). Subjects in the MOD branch tend to have a phenotype that has been associated previously with airway-predominant COPD, with the highest average BMI and lowest amount of emphysema of all branches, as well as thick airways.

Individuals in the SYMPT branch are similar in most aspects to NORM1 and NORM2 branches (mild airway obstruction, low percent emphysema, infrequent exacerbations), yet this group has a larger proportion of current smokers and airway inflammation symptoms, such as cough, phlegm, and chronic bronchitis.

We examined the relation between the five branches and other previously proposed COPD subtypes. As emphysema is a common feature of COPD, recent works have distinguished classified phenotypes as emphysema-predominant (EPD, defined in individuals with GOLD>1 as CT-quantified densitometric emphysema ≥10% at -950 Hounsfield units), non-emphysema-predominant (NEPD, CT emph. <5%) and intermediate emphysema (IE, CT emph. between 5% and 10%) [44]. The breakdown of each branch into separate classes shows that the SEV branch contains the largest fraction of EPD individuals (46%, see Supplementary Figure 3), a smaller fraction of NEPD individuals (23%), and negligible components of other states. Comparatively, the MOD branch is composed of a substantial proportion of NEPD individuals (22% overall, 62% within GOLD>1 stages) and only 8% of EPD phenotypes. Furthermore, MOD has the largest proportion (20%) of individuals in preserved FEV_1_/FVC ratio and impaired spirometry (PRISm). The remaining three branches (SYMPT, NORM1, NORM2) are composed mainly of individuals without significant COPD features and therefore contain negligible proportions of subjects with GOLD stage ≥ 2.

### 2.4 The joint subtypes have distinct transcriptomic signatures and pathway activations

We analyzed the transcriptomic differences between the SEV, MOD, and SYMPT branches using the combined NORM1 and NORM2 branches as the reference group (see Methods). Through differential expression (DE) analysis, we identified a set of DE genes for each contrast (see Supplementary Table 4). For each set, we performed gene set enrichment analysis (GSEA) [45] using the 50 hallmark pathways of MSigDB [46] to find the over- or under-expressed biological pathways in each branch. The results are shown in Fig. 3, where it is evident that the SEV and MOD groups differ markedly in the expression of multiple biological pathways. Among the most significant pathways (FDR p-adj.<0.05), Interferon Alpha (IFN-*α*) Response is highly over-expressed in the SEV and SYMPT branches and under-expressed in the MOD branch. The Oxidative Phosphorylation pathway is upregulated in both the SEV and MOD branches, while the Reactive Oxygen Species (ROS) pathway is upregulated only in the SEV branch and downregulated in the MOD branch. The majority of GSEA leading genes in the MOD branch are antioxidant agents. Among these, differential expression analysis reveals downregulation of the antioxidant enzymes GPX3, G6PD, GSR, and TXNRD2 [47, 48].

**Figure 3:**
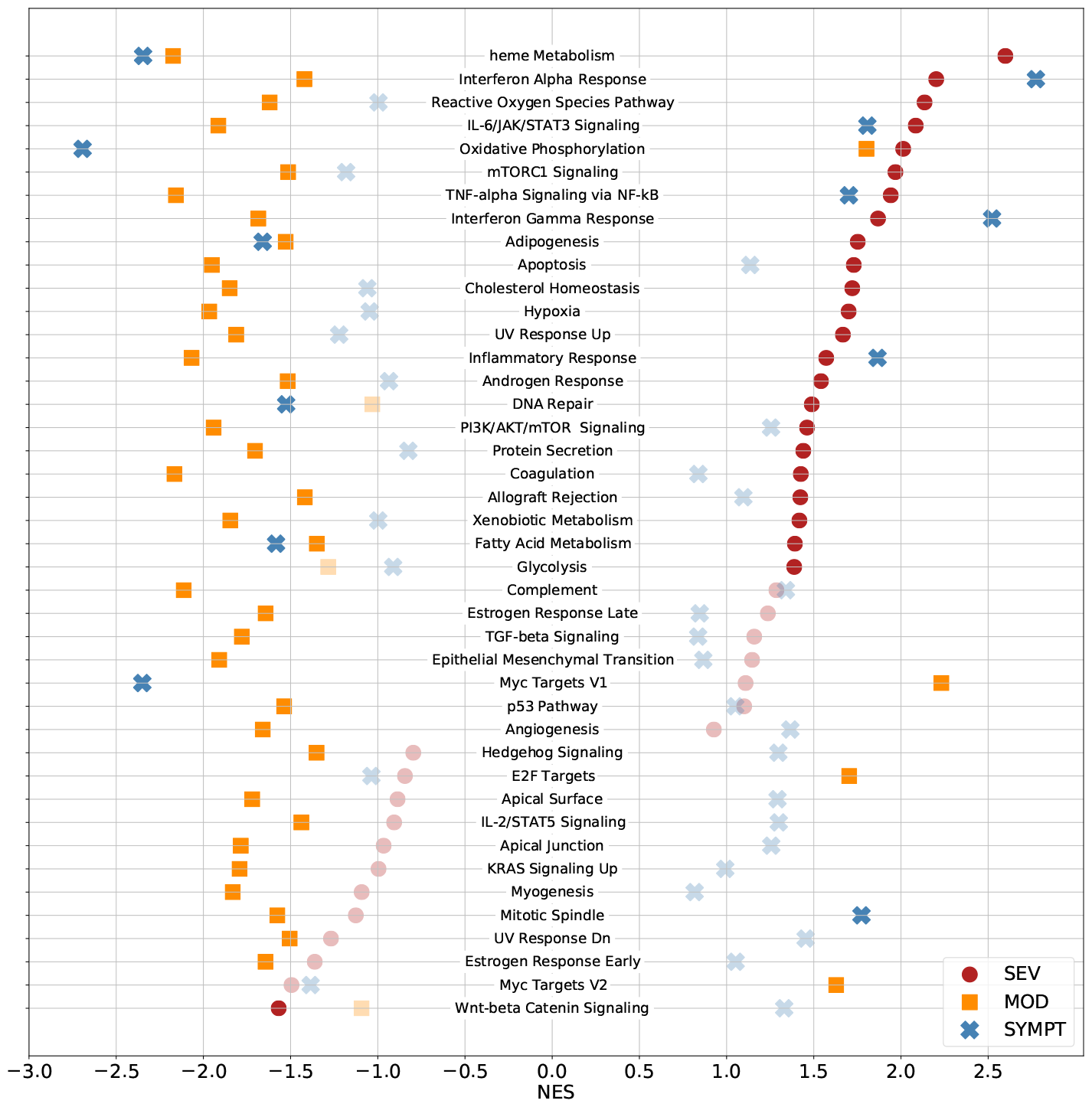
Gene set enrichment analysis of the differentially expressed genes between each branch and the NORM1 and NORM2 branches. Opaque points represent pathways that are significant (FDR p.adj.<0.05), while transparent points are non-significant. NES=normalized enrichment score.

### 2.5 The joint subtypes are associated to distinct disease outcomes and risks

Next, we examined the associations between the joint subtypes and a set of COPD-related clinical outcomes. We collected data from the LFU dataset, in which COPDGene participants self-reported health updates via a survey every 6 months during the whole duration of the study (see Methods). Given that respiratory exacerbations are associated with COPD progression, we examined their temporal patterns among subjects from various branches. The temporal exacerbation patterns of the branches closely mirror the cross-sectional behavior observed during Phase 2 (Fig. 4 (a)), with the individuals in the SEV and MOD branches experiencing a higher rate of exacerbations throughout the entire time period. Interestingly, individuals who reported zero exacerbations in Phase 2, yet were classified in the SEV (163) and MOD (124) branches, demonstrated a significantly higher likelihood of experiencing one or more exacerbation events following Phase 2 — 42% and 50%, respectively — compared to the NORM1 and NORM2 branches (8%) and the SYMPT branch (16%). This finding suggests that branch membership can provide insights into the potential for future exacerbations, even when the present data does not explicitly indicate it. Moreover, to address potential confounding factors, we used a Poisson regression model to analyze the incidence of exacerbations in the LFU data, adjusting for the age, sex, and race of each participant (see Methods). The calculated incidence rate ratios of each branch, using NORM2 as the reference, demonstrate that these insights are consistent even when adjusting for demographic differences between the subpopulations (Fig 4 (b)).

**Figure 4:**
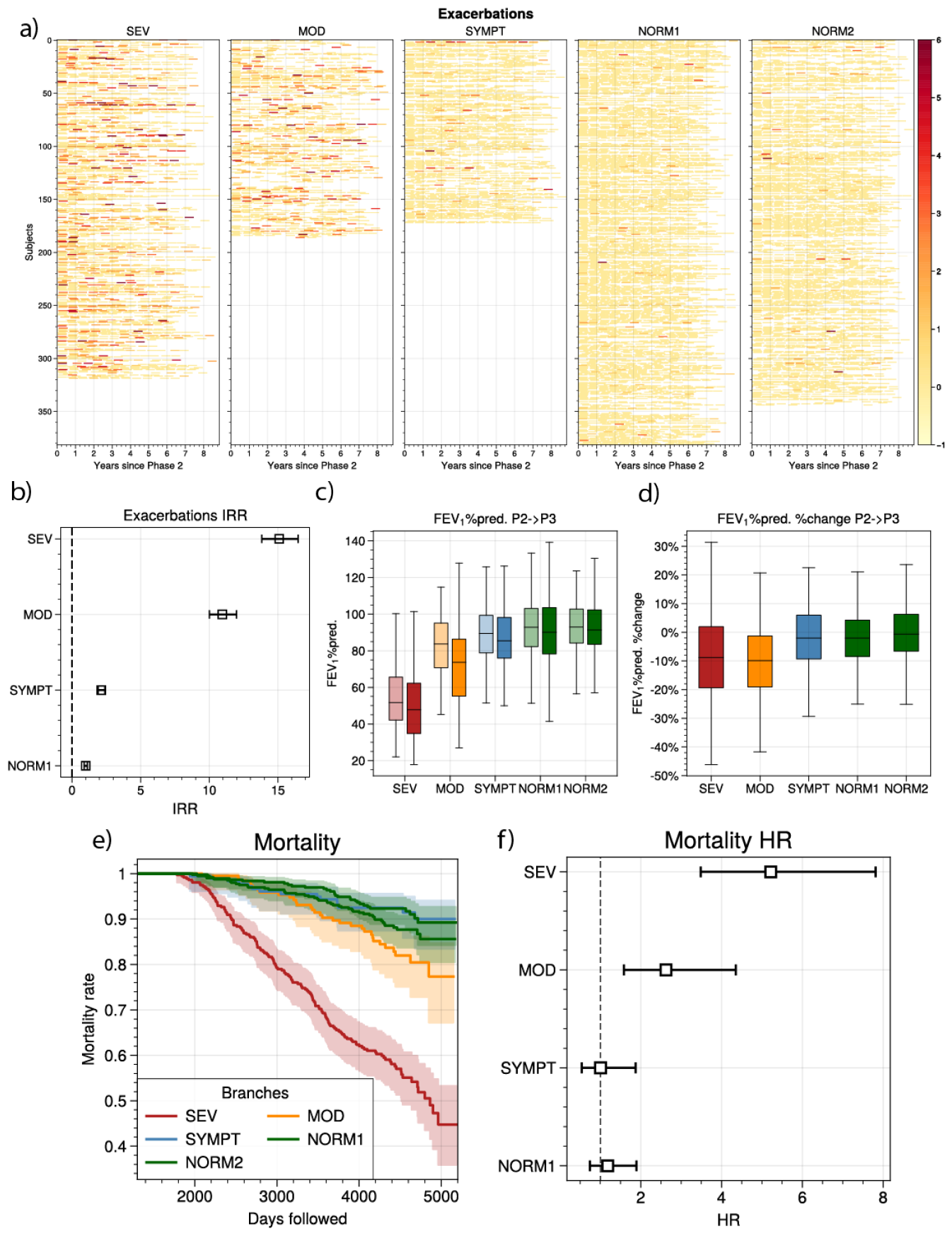
Prospective analysis of the branches. (a) Temporal trends of exacerbations among COPDGene subjects, categorized by their respective branches. Each row represents a subject, and different colored areas within these rows denote the number of exacerbations experienced by that subject during a 6-month timeframe. The color intensity corresponds to the number of exacerbations, with darker or larger areas indicating a higher number of exacerbations. (b) Incidence rate ratio (IRR) of exacerbations between each branch and the reference group, NORM2. (c) Distribution of FEV_1_ % of predicted levels categorized by branch (color), and by study phase (light=P2, dark=P3). (d) Distribution of relative changes in FEV_1_ % of predicted categorized by branch. (e) Kaplan Meier curves of mortality, categorized by branch. (f) Hazard ratio (HR) of mortality between each branch and the reference group, NORM2.

Next, we assessed the change in FEV1 % of predicted values over the five years between the Phase 2 and Phase 3 visits (Figs 4 (c) and (d)). The SEV and MOD branches showed the largest decline in FEV_1_ % of predicted, whereas the other three branches had a smaller decrease within the selected time frame. Finally, we estimated the mortality risk associated with each branch(see Fig. 4 (e) and (f) and Methods). The SEV and MOD branches demonstrate respectively a 5-fold and 2-fold average hazard ratio compared to the NORM2 branch, while the other branches do not exhibit increased risk.

### 2.6 The joint subtypes are robust to retraining and data resampling

To quantify the stability of the VAE space with respect to resampling of the training data, we performed two different robustness tests. In the first test, we evaluated the stability of the PIPs to random re-samplings of the training data. We retrained the VAE 100 times with different train/test splits (80%/20%, selected randomly) and generated 100 new sets of embeddings of the whole population (“resampled” embeddings). Each of these sets of embeddings correspond to the PIPs that would be generated by the VAE under scenarios where different subsets of the data were to be held out. In the ideal case, the resampled embeddings should provide similar information as the original embeddings, indicating that the identified patterns of variability are general and robust to noise. Therefore, we measured the overall similarity between the original and each resampled set of embeddings using the distance correlation measure (dCorr) [49]. dCorr is an extension of the Pearson correlation to multivariate settings and it ranges between 0, indicating statistical independence, and 1, suggesting a linear relationship between the two variables (see Methods for further details). We measured a distribution of correlation values of 0.900±0.006, indicating a strong similarity between the generated profiles.

As a second test, we tested the stability of the branch assignments to random resamplings of the training data. For each of the resampled embeddings described above, we constructed a principal graph (using the same settings) and assigned each point to a branch of the new graph (resampled branches). To measure the robustness of each of the original branches (SEV, MOD, SYMPT, NORM1, and NORM2), we defined the cluster purity measure as the proportion of subjects of each branch that were classified within the same branch in a resampled graph (see Methods). A high cluster purity indicates that individuals within the same cluster tend to be classified in the same cluster also in the resampled configurations. For each branch, we evaluated its purity against each resampled branch classification. In this way we obtained a set of 100 purity values per branch. For comparison, we repeated a similar operation by running the k-means algorithm on both the original VAE embeddings and each resampled embedding. For each embedding, we set a number of clusters equal to the number of branches identified within that embedding. Finally, we evaluated the purity values of each of the clusters retrieved in the original VAE embedding. Since the selection of non-terminal branches in the trajectory analysis has no equivalent operation in the clustering case, for the comparison we selected the 5 clusters with the highest median purity values. Additionally, to emulate the selection of the 50% highest confidence points along the trajectories, for each cluster we selected the 50% data points that are closest to the cluster centroid (‘core set’ of the cluster). The identified branches yield high purity values, approximately 90% for all branches (Supplementary Figure 1 (g), red boxes), outperforming both the standard k-means clusters (light blue boxes) and their core sets (dark blue boxes). This finding indicates that the branches are more robust to random resampling compared to the k-means clusters, suggesting that the principal graph description better reflects the continuous nature of the underlying data distribution.

## 3 Discussion

In this study, we proposed a novel approach to COPD subtyping, bridging the often-separate realms of clinical phenotyping and omics-driven profiling. Our main contributions are: (1) we showed that by integrating clinical and molecular data through variational autoencoders we retain domain-specific information while simultaneously capturing variability across multiple domains; (2) we showed that trajectory analysis in the joint clinical and molecular space of COPD features identifies more robust subgroups compared to standard clustering approaches; (3) we identified 5 joint subtypes with distinctive clinical and transcriptomic features and disease outcomes.

The rationale of this work is that when clinical features aren’t explicitly included in the discovery phase of subtyping, it can be challenging to ensure that the derived molecular subtypes hold clinical relevance [10]. Adding to previous work in multi-omic integration [19, 50], we designed a variational autoencoder architecture to integrate blood gene expression and clinical variables in COPD. While previous studies have focused on finding associations between clinical and molecular variables in specific contexts, such as CT imaging data [51], our integrative methodology describes this connection at a larger scale. Instead of focusing on a specific feature domain, we generated a comprehensive description of the COPD heterogeneity across multiple domains, including transcriptomics, demographics, lung function, lifestyle, CT measures, medical history, comorbidities, and symptoms.

Linear multi-omics integration methods, such as MOFA [22], assume a linear relationship among the features. In contrast, variational autoencoders offer flexibility in capturing complex, non linear interactions between features coming from different domains. The VAE architecture at the core of our methodology is designed to perform an implicit 2-step integration process. The first step consists in integrating features within the same domain (clinical or molecular). Then, the domain-specific higher-level features are subsequently integrated together to encode the Personalized Integrated Profile of a subject.

Several recent works have suggested that COPD manifestations are usually distributed as a continuum rather than discrete subgroups [18, 3], possibly stemming from the superposition of multiple endotypes [52]. In the absence of clearly-separated subpopulations, subjects with intermediate or hybrid COPD conditions are only loosely associated to their subtypes, and even slight noise perturbations can cause these subject to “switch” to adjacent groups. To address this issue, we used the elPiGraph trajectory learning algorithm [36] to map the main trajectories of the VAE latent space. Trajectory analysis yields an explicit linear ordering of individuals along each branch, generalizing the previously proposed concepts of “treatable traits” [52] and “disease axes” [53, 3]. We selected subjects with more extreme conditions to define subgroups, reducing the susceptibility to group switching. This approach improves the subtypes’ robustness compared to standard clustering-based classifications, since in the latter the points with the highest confidence are those nearest to the centroid, and therefore reflect more average, rather than extreme, features.

Our analysis identified five joint subtypes with distinct phenotypic characteristics, severity, disease outcomes, and transcriptomics signatures. Three of these five subtypes present multiple COPD-like features. The subtype with the most severe COPD manifestations, SEV, includes the largest fraction of individuals with emphysema. The SEV group has the largest overlap with the established definition of emphysema-predominant phenotypes (>10% CT-quantified emphysema), a subtype previously associated to larger annual FEV_1_ loss and higher risk of mortality [44]. Conversely, MOD individuals with moderate-to-severe COPD (GOLD≥2) have large overlap with the non-emphysema-predominant subtype of COPD (<5% CT-quantified emphysema). Individuals within the MOD subtype, despite their relatively younger average age and lower disease severity, experience frequent exacerbations and lose lung function at a comparable rate to the SEV group, consistent with prior observations linking exacerbations to loss of lung function [54, 55, 56, 57]. Notably, the fraction of frequent exacerbators in MOD (≥2 exac. per year, as defined in [42]) is composed predominantly (83%) of mildly obstructed individuals (GOLD≤2 or PRISm). The association between frequent exacerbations and milder airflow obstruction in certain COPD subgroups has been demonstrated previously, both within COPDGene (Phase 1) [54] and within the ECLIPSE study [42]. Individuals in the MOD subtype may be representative of earlier-stage COPD mechanisms where frequent exacerbations precede the development of severe airflow obstruction. An alternative possibility is that they represent a distinct trajectory of COPD characterized by airway inflammation and frequent exacerbations without emphysematous lung destruction.

Subjects in the SYMPT subtype are mostly current smokers (∼70%) with only slight spirometric abnormalities and relatively favorable disease progression, yet they suffer from multiple respiratory symptoms, including chronic bronchitis, coughing and wheezing. SYMPT individuals are reminiscent of the previously investigated subtype of symptomatic smokers with preserved lung function (FEV_1_:FVC≥0.70) [58, 59]. In agreement with the findings in [58], using LFU measurements we found that subjects within this subtype experience higher exacerbation frequency compared to the NORM2 baseline. However, in our prospective analysis we did not observe a substantial difference in loss of lung function between the SYMPT and the NORM1 and NORM2 subtypes.

Besides recapitulating previously observed patterns of phenotypic variation, the joint subtypes display marked transcriptomic differences. We measured a distinct pattern where multiple biological pathways exhibit an opposite directionality of activation between the SEV and MOD branches. Immune pathways, which are among the top enriched processes, have been consistently associated to COPD [11, 60, 61]. Among the most significant differences, we found several pathways related to immune response and regulation, including Interferon Alpha response, IL6/JAK/STAT3 Signaling and TNF-*α* Signaling via NF-*κ*B. COPD patients who experience frequent exacerbations have been reported to exhibit reduced IFN-*α* levels in response to viral infection compared to individuals with lower exacerbation rates [62]. As such, downregulation of the IFN-*α* in the MOD branch might indicate compromised antiviral immunity, potentially leading to a higher susceptibility to exacerbations. ROS overproduction is known to suppress the activity of these enzymes [63, 64, 65, 66], and prolonged depletion of antioxidant capacity has been observed several days after the onset of exacerbation in COPD patients. Furthermore, previous reports have supported the role of the IL6/JAK/STAT3 signaling pathway in pulmonary inflammation and COPD severity [67, 68].

One limitation of this study is that we rely exclusively on expression data in blood for inferring the molecular processes associated to each subtype. The integration of diverse omics types and tissues [19], especially from the lung and airways, will be crucial for delineating more detailed disease subtypes that capture the diversity of COPD processes and their relationship with its clinical manifestations. Another limitation of our study is the lack of replication of our results in independent cohorts. Independent validation of this analysis is challenging since there are currently no other cohorts that have collected the data required in enough subjects with advanced COPD. In the future, with the additional generation of multi-omics data in NIH-funded studies such as SPIROMICS [69] through the Trans-Omics in Precision Medicine (TOPMed) program [70], it will be possible to pursue independent validation. In the meantime, we have pursued the next-best option, namely an extensive robustness analysis using resampling approaches. A crucial objective left for future work is the generation of distilled models that can reproduce our results in reduced datasets. Finally, cell type proportions play a dual role in gene expression analyses, serving both as a potential manifestation of the disease and a potential confounder [71]. This dual nature hinders the interpretability of gene expression results. Leveraging single cell RNA-seq data offers a way forward [72, 73]. By providing a higher resolution view of individual cell populations and their associated gene expression patterns, it becomes possible to discern between disease-associated shifts in cell populations and gene expression changes within specific cell types. Therefore, another promising future direction for this work is its application to single cell RNA-seq datasets.

## 4 Methods

### 4.1 Processing of clinical and phenotypic data

In the initial phase of preparing the input data for the VAE, we chose all the subjects who had available clinical data and RNA-seq expression profiles, obtaining 3,628 samples. We classified each feature as either numerical or categorical, and we devised different processing strategies to handle the two groups of features. Among the categorical features, we excluded those with more than 10% of missing values across all subjects or where the most frequent category was present in more than 80% of subjects. The latter criterion was devised to avoid including features that are not sufficiently informative of patient heterogeneity. Similarly, we excluded the numerical features with more than 10% missing values or with constant values. We then imputed the remaining missing values in the categorical features by considering the most frequent category for each feature across all subjects. The numerical missing values were imputed through KNN imputation with k equal to 10. As further selection, we selected one representative variable among the groups of redundant categorical variables, i.e., those with high similariy values (adjusted Rand score > 0.95). The resulting set of clinical features selected for training the VAE is summarized in Supplementary Table 1.

### 4.2 Processing of expression data and differential expression analysis

From the raw read counts matrix we removed low expressed transcripts by selecting transcripts with at least 1 CPM in more than 10 samples. Next, we processed the data with the DeSeq2 algorithm [74], and removed batch effects with the function “removeBatchEffect” of the package limma [75]. Finally, to make the computations more manageable, and to perform a preliminary feature selection, we selected only the genes that were loosely associated to at least one clinical feature in the dataset. We assessed the significance of the relationships between each gene and each clinical feature, using Spearman’s correlation for numerical features and the Kruskal Wallis test for categorical ones. Genes with at least one FDR-adjusted p-value below 0.001 were retained, yielding 5,979 transcripts.

To perform differential expression (DE) analysis we executed the DE pipeline of DeSeq2 starting from the raw data. In brief, after basic data filtering we set up a design matrix with covariates including sequencing batch, age, sex, race, and white blood cell proportions. As contrasts we choose the membership to each of the three COPD branches (SEV, MOD, SYMPT) against the joined population of the reference branches NORM1 and NORM2. From the three contrasts we obtained three DE summary statistics. Next, we performed Gene Set Enrichment Analysis [76] with the GSEApy python package [77]. Specifically, we ranked the genes by their negative log p-value score multiplied by the sign of their log fold change (logFC). In this way, the genes that are significantly upregulated in the contrast (low p-value, positive logFC) appear at the top of the ranking, while the genes that are downregulated (low p-value, negative logFC) will appear at the bottom, providing a coarse measure of their general state of differential activation.

### 4.3 Conditional Variational Autoencoder design

In order to find a shared latent space for expression and clinical features, we designed a conditional VAE architecture with an “X” shape, similar to the X-VAE model described in [29], shown in Fig. 1 (d). The network takes as input a vector of concatenated RNA-seq read counts and clinical features that have been normalized to be in the unit range.

The architecture consists of four subnetworks: two encoders 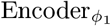 and 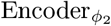, and two decoders 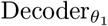 and 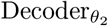. Each data mode *x*_1_ and *x*_2_, along with the conditional variable *c*, is separately passed through an encoder network.

Each encoder network, 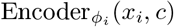 is composed of L layers where each layer *l* is defined as:

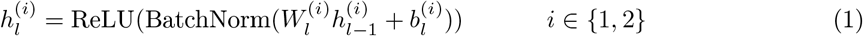

with

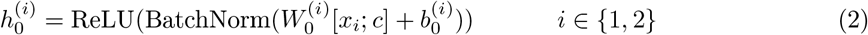

Here, 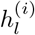 denotes the *l*-th layer’s activations, BatchNorm is a batch normalization transformation, 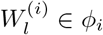 and 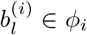 are the weight matrix and bias terms of the *l*-th layer, ReLU is the activation function, and [·;·] is the concatenation operator.

These encoders transform their inputs into latent representations:

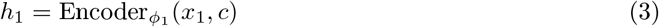

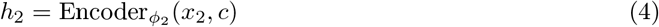

The two latent representations *h*_1_ and *h*_2_ are then concatenated to form a shared representation *h* = [*h*_1_; *h*_2_]. Two separate linear layers are used to project this representation into the parameters of a Gaussian distribution, forming the mean vector *μ*(*h*) and the vector of standard deviations *σ*(*h*) of the latent distribution of the latent vector *z*.

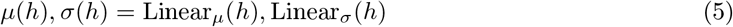

For the decoder part, the sampled latent variable *z* is concatenated with the conditional variable *c* and passed through the decoder networks to generate the reconstructed data 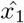 and 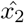.

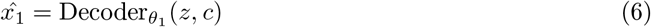

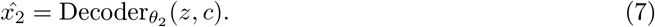

Similarly, a decoder 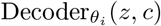 is composed of *L* layers

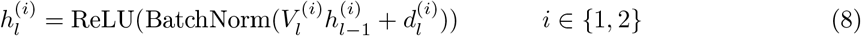

with

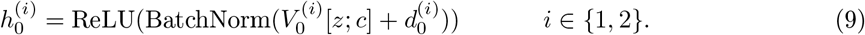

As before, 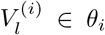 and 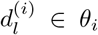 are the weight matrix and bias terms of the *l*-th layer. Furthermore, the last layers of each decoder do not include a ReLU activation function to allow for negative outputs.

#### 4.3.1 The Evidence Lower Bound and Maximum Mean Discrepancy Loss

The training objective of our multimodal VAE is to maximize the Evidence Lower Bound (ELBO), which in this case is a convex combination of the data-type-specific ELBO terms, governed by a parameter *α*_1_. The reconstruction loss for each mode is calculated differently due to the nature of their data. For the numerical data mode *x*_1_, the Mean Squared Error (MSE) is used. For the mixed data mode *x*_2_, which contains numerical and categorical data, the loss is a convex combination (controlled by a parameter *α*_2_) of the MSE for the numerical part and the Categorical Cross-Entropy for the categorical part. Furthermore, instead of the standard Kullback-Leibler (KL) divergence used in the original formulation of VAE [35], we employ the Maximum Mean Discrepancy (MMD) measure to regularize the model as proposed in the InfoVAE formulation [78]. Similarly to KL divergence, MMD is a distance measure between two probability distributions. However, MMD only depends on a set of statistics evaluated from the two distributions, and therefore it does not require the explicit evaluation of their analytical form. In the Gaussian case, MMD calculation is done by embedding the distributions in a Reproducing Kernel Hilbert Space (RKHS) identified by their average and standard deviation, and by computing the distance between these statistics [79]. The ELBO _MMD_ has the form

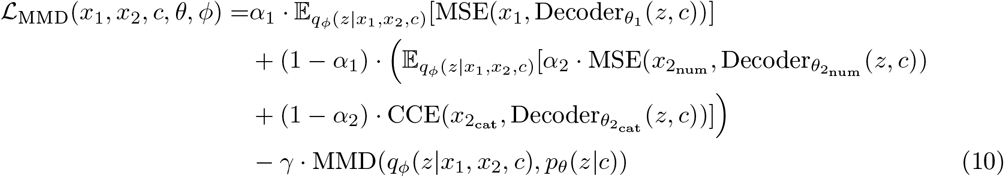

Here, 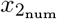 represents the numerical part and 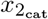 represents the categorical part of the second data mode *x*_2_. The expected values 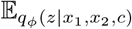 are approximated by a single sampling of *z* ∼ 𝒩 (*μ*(*h*), *σ*(*h*)). 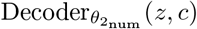 and 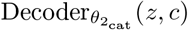 are the reconstructed numerical and categorical parts of the data respectively, and CCE denotes the average Categorical Cross-Entropy loss across all the categorical variables. MMD(*q*_*ϕ*_(*z*|*x*), *p*_*θ*_(*z*)) measures the dissimilarity between the approximate posterior *q*_*ϕ*_(*z*|*x*) and the prior *p*_*θ*_(*z*) in terms of their mean embeddings in the RKHS.

#### 4.3.2 Optimization

Optimization was performed with the ADAM optimizer [80]. In all our tests the full dataset is split in 80% training samples, 10% validation samples and 10% test samples. To identify the optimal hyperparameters (number and size of hidden layers, the learning rate, and training batch size), we performed hyperparameter optimization with ASHA (Asynchronous Successive Halving Algorithm), a scheme for parallel optimization equipped with greedy early stopping strategies to rule out inefficient hyperparameter configurations [81]. In the final optimal configuration the encoder subnetwork for processing the expression data has 2 hidden layers of dimensions 1024 and 512 neurons, while the encoder subnetworks for processing the clinical data has 1 hidden layer of dimension 64 neurons. The decoder networks are constrained to mirror the encoder structure in reverse. The mini-batch size is 128.

### 4.4 Prediction of future outcomes from PIPs

The prediction task was performed by training a set of random forest (RF) classifiers using multiple input embeddings and output disease outcomes, as listed in the main text.

The embeddings of MOFA [22] were calculated by running the mofapy python package, feeding the expression and clinical data as input and by setting 30 as the number of hidden factors. The MOFA embeddings were defined as the estimated means of the factors obtained after fitting the model.

For ease of comparison of performances, we considered a set of binary outcomes. COPD outcomes that are measured as continuous variables were transformed to binary variables by thresholding. The selection of thresholds was guided by practical considerations to ensure a balanced representation of positive and negative examples. The considered target variables are the following: (1) ΔFEV_1_ % of pred. (P3<P2): subjects with more than 10% decrease of FEV_1_ percent predicted between Phase 2 and Phase 3 (positive class) compared to subjects with more than 10% increase (negative class) (*N* =331); (2) inc. bronchitis (P3): incident chronic bronchitis in Phase 3, restricted to individuals without chronic bronchitis in Phase 2 (*N* =1,087); (3) exacerbations (P3): frequency of exacerbations in Phase 3 greater than 0 (*N* =1,251); (4) Δ exacerbation frequency (P3>P2): frequency of exacerbations in Phase 3 greater than Phase 2 (*N* =1,251); (5) severe exacerbations (P3): presence of severe exacerbations in Phase 3 (*N* =1,250); (6) Δ severe exacerbations (P3>P2): presence of severe exacerbations in Phase 3 in subjects who did not experience severe exacerbations in Phase 2 (*N* =1,164); (7) ΔmMRC (P3>P2): increased mMRC dyspnea score in Phase 3 compared to Phase 2 (*N* =1,183); (8) ΔSF-36 (P3<P2): decrease of SF-36 score between Phase 2 and Phase 3 (*N* =1,251); (9,10) mortality (3/5yr): all-cause mortality at 3 and 5 years (*N* =3,361/3,347). We set up each RF classifier with 100 decision trees. For a more robust performance assessment that is not too sensitive to a specific train/test split of the dataset, we conducted a stratified 5-fold cross-validation, repeated 3 times. For each split, we performed three steps: (1) we normalized the whole dataset according to the statistics obtained from the current training set; (2) since most clinical outcomes have highly unbalanced class distributions, we performed SMOTE oversampling [82] of the minority class, using the *imblearn* python package [83]; (3) we trained the classifier with the resulting data. The values shown in Table 1 are the summary statistics obtained by the 5-fold splits repeated 3 times, for a total of 15 performance values for each prediction. The significance values are obtained by performing t-tests between the performance values obtained by the best embedding and the second-best embedding.

### 4.5 Construction of principal graph

To build the principal graph describing the distance relationships between observations in the embedding space, we used the elPiGraph method [36]. We used default parameters, except for the maximum number of nodes which was set to 30 to increase resolution. Also we set the “collapse” argument to True, in order to merge the small and noisy branches within their main branch. All the 2D embeddings shown in Fig. 2 are evaluated with a modified version of the elPiGraph function “visualize_eltree_with_data”. In brief, this function first produces a 2D embedding of the principal graph using the Kamada-Kawai layout algorithm [84]. Next, it distributes all the data points across the branches according to their calculated projections. Finally, to improve clarity each point is scattered randomly in the direction orthogonal to the branch by an extent controlled by a fixed parameter.

### 4.6 Processing of LFU and mortality data

To visualize the trends in Fig. 4 (a), we considered all the long-term follow-up (LFU) survey data that were compiled after Phase 2 of the COPDGene study (as of August 2022). Since the time points refer to the time the survey was compiled, we considered as the interval range of each data point the 6 months prior to the compile date, unless another survey was compiled by the same subject less than 6 months earlier. In that case, the time interval is the time span occurring between the two surveys. To analyze the risk of increased exacerbations over time, we set up a Poisson regression model, controlling for the age, sex, and race covariates. The model was fit through the *glmfit* R function, using a log link function. We also tested an alternative mixed effect model where subject identity was included as a random effect, obtaining similar results. To estimate mortality at 3 and 5 years, we considered the COPDGene all-cause mortality data as of October 2022. To implement the Cox proportional hazard model of mortality we used the *lifelines* python package [85].

### 4.7 Evaluation of distance correlation (dCorr)

The similarity between two sets of *N* vectors embedded in two spaces can be estimated by modeling each vector set as the *N* realizations of a multivariate random variable. From this standpoint, the similarity between the two sets is equivalent to the level of statistical dependency between the two variables. dCorr is an extension of the Pearson correlation to multivariate settings and it ranges between 0 (statistical independence) and 1 (linear dependency) [49]. Furthermore, dCorr is invariant to rigid transformations applied to either of the two spaces (e.g. rotations). This makes it an ideal tool for assessing the similarity between the two sets of vector embeddings. dCorr is estimated as follows [86]:

let *X* and *Y* be two *d*-dimensional vector sets. Define *a*_*ij*_ and *b*_*ij*_ to be the Euclidean distances between the *i*th and *j*th elements of *X* and *Y*, respectively. We then form the centered distance matrices *A* and *B* as follows:

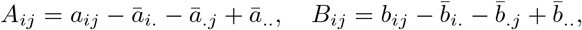

where

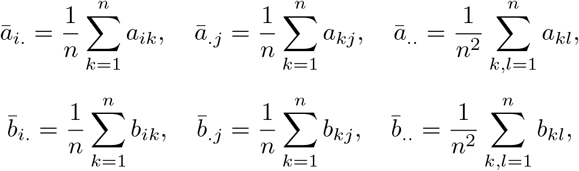

The distance covariance (dCov) and distance correlation (dCorr) are defined as

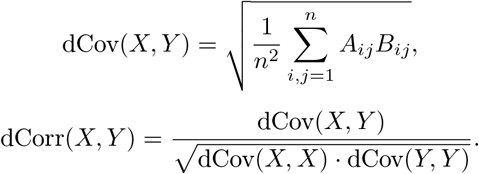

The distance covariance correlation was evaluated with the python package *hyppo* [87].

### 4.8 Evaluation of the branch purity with respect to data resamplings

Let us consider a set of *n* data points with branch labels *Y* = {*y*_1_, *y*_2_, …, *y*_*n*_}, where each *y*_*i*_ belongs to one of *K* branches. In a resampled embedding we produce a new branch assignment 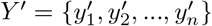, where each 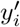 belongs to one of *K*^*′*^ branches, with *K* not necessarily equal to *K*. The purity of branch *k* in the original labeling *Y* with respect to the resampled branch labels *Y* is defined as

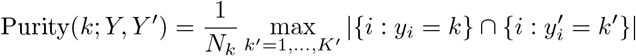

where *N*_*k*_ = |{*i* : *y*_*i*_ = *k*}| and |. | denotes the cardinality of a set. This value measures the number of items in the *k*-th branch that belong to the most common resampled branch. Purity is a fraction between 0 and 1, where higher values indicate stronger alignment between the original and resampled branch assignments.

## Data Availability

All data used in this study are available in dbGaP phs000179.v6.p2

## 5 Data Availability

All COPDGene data for the analysis are available in dbGaP accession number phs000179.v6.p2. The data generated in this study has been deposited in the Zenodo repository, accessible via the DOI: 10.5281/zenodo.10431493.

## 6 Code Availability

The code used to reproduce the analyses in this study is available in the GitHub repository accessible via the following URL: https://github.com/reemagit/joint_subtyping_vae.

## 7 Author contributions

EM, PJC and KG conceptualized the study and designed the project. EM led the data analysis and code implementation. MDM, ZX, RTC, and JHY contributed to the data processing steps. MDM, PJC, and KG contributed to the data analysis. CPH, STW, EKS, PJC provided domain expertise for the analysis and interpretation of the results. EM was the lead author of the manuscript. All authors contributed to writing and revising the manuscript.

## 9 Acknowledgments

This work was supported by grants from the NHLBI (EM: K01HL166705; MDM: K25HL168157; STW: PO1HL132825; KG: R01HL155749). The COPDGene study (NCT00608764) is supported by grants from the NHLBI (U01HL089897 and U01HL089856) and by NIH contract 75N92023D00011 and by the COPD Foundation through contributions made to an Industry Advisory Committee that has included AstraZeneca, Bayer Pharmaceuticals, Boehringer-Ingelheim, Genentech, GlaxoSmithKline, Novartis, Pfizer and Sunovion.

